# Chronic lithium therapy and urine concentrating ability in individuals with bipolar disorder: association between daily dose and resistance to vasopressin and polyuria

**DOI:** 10.1101/2022.01.28.22270045

**Authors:** Nahid Tabibzadeh, Emmanuelle Vidal-Petiot, Lynda Cheddani, Jean-Philippe Haymann, Guillaume Lefevre, Bruno Etain, Frank Bellivier, Emeline Marlinge, Marine Delavest, François Vrtovsnik, Martin Flamant

## Abstract

**Background and objectives:** Chronic lithium treatment in individuals with bipolar disorder can induce nephrogenic diabetes insipidus. However, the prevalence, kinetics and mechanisms of such complication are poorly known. We aimed at evaluating patterns of urine concentrating ability and the correlates of 24-hour urine output in individuals treated with lithium.

**Design, setting, participants and measurements:** Prospective single center observational study of 217 consecutive individuals treated with lithium carbonate and referred to the renal unit. All individuals collected 24-hour urine the day before admission and underwent a desmopressin (DDAVP) concentrating test, fasting plasma vasopressin measurement (copeptin measurement in a subset of individuals, n=119), and measured GFR (mGFR) using urinary ^99^Tc-DTPA clearance. Maximal urine osmolality (Max Uosm) was defined as the highest level during the DDAVP test.

**Results:** 21% of individuals displayed polyuria (> 3l/day), but 55% displayed elevated fasting vasopressin level (> 5 pg/ml). During the DDAVP test, Uosm was significantly lower, and urinary output and free water clearance were significantly higher in the highest treatment duration tertile (> 10 years) whereas no difference was observed between the first two tertiles (< 2.5 years and 2.5-10 years). Among individuals with normal Max Uosm (>600 mOsm/KgH_2_O) (n=128), 51% displayed elevated vasopressin levels, which was associated with higher lithium daily doses (950 [750- 1200] versus 800 [500- 1000] mg/d, p<0.001), and 100% of patients with lithium daily dose ≥1400 mg/d had high vasopressin levels. In multivariable analysis, 24-hour urine output was associated with higher lithium daily dose (β 0.49 ± 0.17, p=0.005), female sex (β -359 ± 123, p=0.004), daily osmolar intake (β 2.21 ± 0.24, p<0.001), maximal urine osmolality (β -2.89 ± 0.35, p<0.001) and plasma vasopressin level (β 10.17 ± 4.76, p=0.03), but not with lithium formulation.

**Conclusions:** Higher lithium daily dose was associated with higher vasopressin levels and higher urine output, independently of other factors. Daily osmolar intake was also associated with higher 24-hour urine output. These results suggest that controlled salt and protein intake and lithium dose might reduce renal resistance to vasopressin in these patients.

## Introduction

Lithium carbonate represents the cornerstone treatment of bipolar disorder. Its effectiveness to reduce the risk of maniac or depressive relapse has been demonstrated in several studies (1–3). In addition, lithium has demonstrated its efficacy in suicide prevention (4). This efficacy is however balanced by short and long-term side effects including renal and metabolic complications (5). One of the most frequent renal side effects is nephrogenic diabetes insipidus (NDI), resulting in polydipsia and polyuria (6,7). NDI has been described in humans as well as in experimental animal models and develops shortly after the initiation of treatment. In healthy volunteers, NDI appears 8 days after lithium treatment initiation, and resolves shortly after discontinuation (8). In previous studies, 20 to 70% of lithium-treated individuals had NDI depending on the population and on the definition of NDI (9).

The mechanism of lithium-induced NDI is partially known. It implicates the down-regulation of type 2 aquaporins (AQP2) expression and addressing to the apical membrane of the principal cells in the collecting duct of the nephron (10). How lithium induces this decrease in AQP2 expression is still a matter of debate. In individuals treated with lithium, risk factors for developing lithium-induced polyuria are still poorly known. We thus aimed at evaluating the concentrating ability and determining the factors associated with a decrease in urine concentrating ability in a cohort of 217 individuals treated with lithium.

## Methods

### Study design and population

From March 2015 to December 2020, 230 consecutive adult patients receiving lithium for a bipolar disorder were referred for a systematic assessment to the Department of Renal Physiology by psychiatrists and nephrologists. Among these patients, we excluded 13 individuals who had discontinued lithium treatment at the time of admission, leaving 217 individuals. Eligible patients were ≥ 18 years of age at inclusion, with all durations of lithium treatment, and had neither started dialysis nor underwent kidney transplantation.

The local ethics committee approved the study (APHP.Nord Institutional Review Board CER-2021-74). All individuals provided written informed consent before inclusion in the study cohort.

### Data collection and measurements

During a 5-hour in-person visit, a large set of clinical and laboratory data were collected including GFR measurement using renal clearance of a radioisotope, as previously described (11). Individuals were instructed to fast (not to eat or drink) from 8 p.m. the day before the admission. Individuals were asked to collect their 24-hour urine the day before admission. A written information document was provided and indications to discard the first morning void on the first day, and then to collect all urine including the first void the next morning were given by a trained nurse. In addition to this information, 24-hour urine collection was verified by the interrogation of the patient, and corrected using the fractionated creatinine clearance during the test (12). A kidney MRI was performed in a subset of patients (n=108). Along with these tests, a urine two-step concentrating test was performed. First, fasting urine and plasma osmolality were measured on the first-morning urine sample, along with natremia, plasma copeptin level (B·R·A·H·M·S™ Copeptin proAVP KRYPTOR™, 1.7 < normal value < 11.25 pmol/l, performed in a subset of 119 individuals), plasma vasopressin level (in-house radioimmunology assay, 1< normal value < 5 pg/ml, Tenon Hospital Laboratories) as previously described (13). Urine output, ionogram, urea, creatinine and osmolality were determined on 24-hour urine collection. 24-hour osmolar intake was defined by the 24-hour urinary osmolar output. Thirst score was determined at each time point by trained nurses using a numerical scale from 0 (not thirsty at all) to 10 (the worst thirst imaginable). A challenging test with desmopressin (1-deamino-arginine vasopressin, DDAVP) was performed, which was injected 2 hours after the admission of the patient who remained fasting. Briefly, urine and blood samples were collected every 30 minutes over a 4-h period following 4□µg intravenous administration of DDAVP in order to measure natremia, plasma and urine osmolality. Individuals were allowed to eat a breakfast after DDAVP injection, and to drink one glass of water at each time point. The highest value of urine osmolality among all measures was used to define the maximal urinary osmolality. Normal concentrating ability was defined as a maximal urine osmolality above 600 mOsm/kgH_2_O, as previously described (14). Free water clearance (FWC) was calculated as follows:

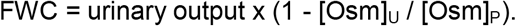

Polyuria was defined as a 24-hour urinary output above 3 liters (15).

### Statistical analyses

Categorical variables were described as numbers and percentage, and continuous variables as mean ± SD or median [Quartile 1 Q1-Quartile 3 Q3], as appropriate. Baseline individuals’ characteristics were compared according to tertiles of lithium treatment duration, and among subgroups according to urine concentration features using analysis of variance (ANOVA) (or Kruskal–Wallis test, as appropriate) and the Chi-square test, for quantitative and qualitative variables, respectively. The 4 subgroups according to urine concentrating features were the following: group 1: normal urine concentrating ability defined as: maximal urine osmolality>600 mOsm/kgH_2_O, and normal vasopressin level, < 5 pg/ml; group 2: partial resistance to vasopressin defined by a vasopressin level>5 pg/ml but normal maximal urine osmolality; group 3: complete resistance to vasopressin defined by a vasopressin level> 5 pg/ml and an impaired urine concentrating ability defined by a maximal urine osmolality< 600 mOsm/ kgH_2_O; group 4: normal vasopressin level < 5 pg/ml and impaired maximal urine concentrating ability < 600 mOsm/ kgH_2_O consistent with primary polydipsia. Simple and multiple linear regression analyses were performed to investigate the determinants of 24-hour urinary output. Models were compared using log-likelihood statistics, to select the statistically better multiple linear regression model. A stepwise variable selection method was used (nested models). The absence of multicollinearity was verified by calculating the Variance Inflation Factor (VIF).

Validity conditions of the multiple linear regression model were checked. First, the model’s suitability was tested using the Rainbow test (16). We then studied the residuals. Their independence was checked using the Durbin-Watson test associated with a graphical method (17). A quantile-quantile plot was made to check their normal distribution. The distribution’s homogeneity has been checked graphically by representing the square root of standardized residuals versus fitted values. Covariates were selected *a priori*, including maximal urinary osmolality, mGFR, 24-hour osmolar intake, fasting thirst score, lithium treatment duration, lithium dose, lithium formulation, sex, ionized calcium, extracellular volume, hypertension (defined as treated hypertension or blood pressure > 140/90 mmHg), plasma vasopressin, age and body mass index.

A two-sided p-value < 0.05 indicated statistical significance. Statistical analyses were performed with the R software, using the R-Studio interface (version 1.2.5019) (18).

## Results

### General description of the population

Individuals’ characteristics in the total population and according to lithium treatment duration tertiles (tertile 1: <2.5 years; 2.5 ≤ tertile 2 <10 years; tertile 3 ≥ 10 years) are reported in table 1. Median age was 51 [37-62] years, 38% of the individuals were male, median mGFR was 78 [63-90] ml/min/1.73m^2^, and median lithium treatment duration was 5 [2-14] years. Median 24-hour urine output was 1932 [1448-2835] ml/day and 21% of the patients displayed polyuria. Median fasting plasma copeptin and vasopressin levels were elevated, respectively 13 [8-23] pmol/l and 5.8 [3.9-10.3] pg/ml. Compared to individuals in the first and second tertile of lithium treatment duration, individuals in the third tertile of lithium treatment duration were significantly older, had a lower daily dose of lithium, a higher 24-hour urine output, a lower maximal urine osmolality, higher copeptin and vasopressin levels, with no difference in 24-hour osmolar intake or plasma lithium level. mGFR was also significantly lower and renal microcysts on MRI were significantly more frequent (table 1).

**Table 1.** Characteristics of the population.

|  |  | Treatment duration |  |  |  |
| --- | --- | --- | --- | --- | --- |
|  | Overall | Tertile 1<br>< 2.5 years | Tertile 2<br>2.5-10 years | Tertile 3<br>> 10 years |  |
| n | 217 | 68 | 70 | 79 | p |
| Age, years | 51 [37-62] | 38 [30-53] | 45 [36-58] | 60 [52-66] | <0.001 |
| Male sex n (%) | 83 (38) | 31 (46) | 25 (36) | 27 (34) | 0.3 |
| Hypertension n (%) | 56 (26) | 20 (29) | 17 (25) | 19 (24) | 0.7 |
| BMI, kg/m<sup>2</sup> | 26 [23-29] | 26 [23-28] | 26 [23-30] | 26 [23-29] | 0.6 |
| Lithium treatment duration, years | 5 [2-14] | 1 [0.5-2] | 4 [3-6] | 18 [13-26] | <0.001 |
| Sustained-release formulation | 139 (65) | 49 (74) | 48 (71) | 42 (53) | 0.02 |
| Lithium daily dose, mg/d | 800 [575-1000] | 1000 [688-1200] | 800 [500-1000] | 750 [500-888] | 0.004 |
| Serum lithemia, mmol/L | 0.7 [0.6-0.9] | 0.8 [0.6-0.9] | 0.7 [0.6-0.8] | 0.7 [0.6-0.9] | 0.8 |
| 24-hour urine output, mL/d | 1932 [1448-2835] | 1662 [1227-2238] | 1765 [1460-2443] | 2516 [1794-3265] | <0.001 |
| Polyuria > 3L/d | 46 (21.4) | 9 (13) | 12 (17) | 25 (32) | 0.01 |
| 24-hour osmolar intake, mOsm/24h | 659 [516-847] | 645 [506-887] | 669 [507-837] | 663 [551-829] | 1 |
| Fasting thirst score, on a 0-10 scale | 5 [4-7] | 6 [4-7] | 5 [4-7] | 5.00 [4.00, 7.00] | 0.3 |
| Fasting natremia, mmol/L | 141 [140-142] | 141 [140-141] | 141 [140-142] | 142 [141-144] | <0.001 |
| Fasting Uosm, mOsm/kgH<sub>2</sub>O | 578 [408-719] | 689 [545-757] | 621 [500-737] | 406 [295-562] | <0.001 |
| Maximal Uosm, mOsm/kgH<sub>2</sub>O | 716 [547-853] | 823 [706-898] | 789 [636-870] | 529 [385-683] | <0.001 |
| Plasma copeptine, pmol/L | 13 [8-23] | 11 [8-16] | 11 [7-14] | 24 [14-42] | <0.001 |
| Plasma vasopressin, pg/mL | 5.8 [3.9-10.3] | 5 [3.3-7.8] | 5.7 [3.9-8.5] | 7.7 [4.6-13.5] | 0.005 |
| mGFR, ml/min/1.73m<sup>2</sup> | 78 [63-90] | 88 [76-96] | 84 [71-94] | 63 [46-77] | <0.001 |
| Renal microcysts | n=99 |  |  |  | <0.001 |
| 0 n (%) | 49 (50) | 19 (76) | 23 (74) | 7 (16) |  |
| 1 to 10 n (%) | 30 (30) | 6 (24) | 8 (26) | 16 (37) |  |
| 11 to 50 n (%) | 10 (10) | 0 (0) | 0 (0) | 10 (23) |  |
| > 50 n (%) | 10 (10) | 0 (0) | 0 (0) | 10 (23) |  |

Data are expressed as median (Q1-Q3) or number (percentage). Comparisons were performed using Kruskal-Wallis test or Chi-squared test. BMI: body mass index; Uosm: urinary osmolality. mGFR: measured glomerular filtration rate.

### DDAVP test according to lithium treatment duration tertiles

Throughout the DDAVP challenging test, individuals in the highest lithium treatment duration tertile (> 10 years), displayed lower mean urine osmolality, higher urine output and higher free water clearance throughout the test, compared to the first two tertiles.

Urinary output decreased significantly 60 minutes after DDAVP injection in all tertiles, and tended to increase afterwards, but with no significant increase in urine osmolality or decrease in free water clearance (figure 1). There was no difference in patterns of urine osmolality, urine output and free water clearance along the test in the first two tertiles of lithium treatment duration.

**Figure 1.**
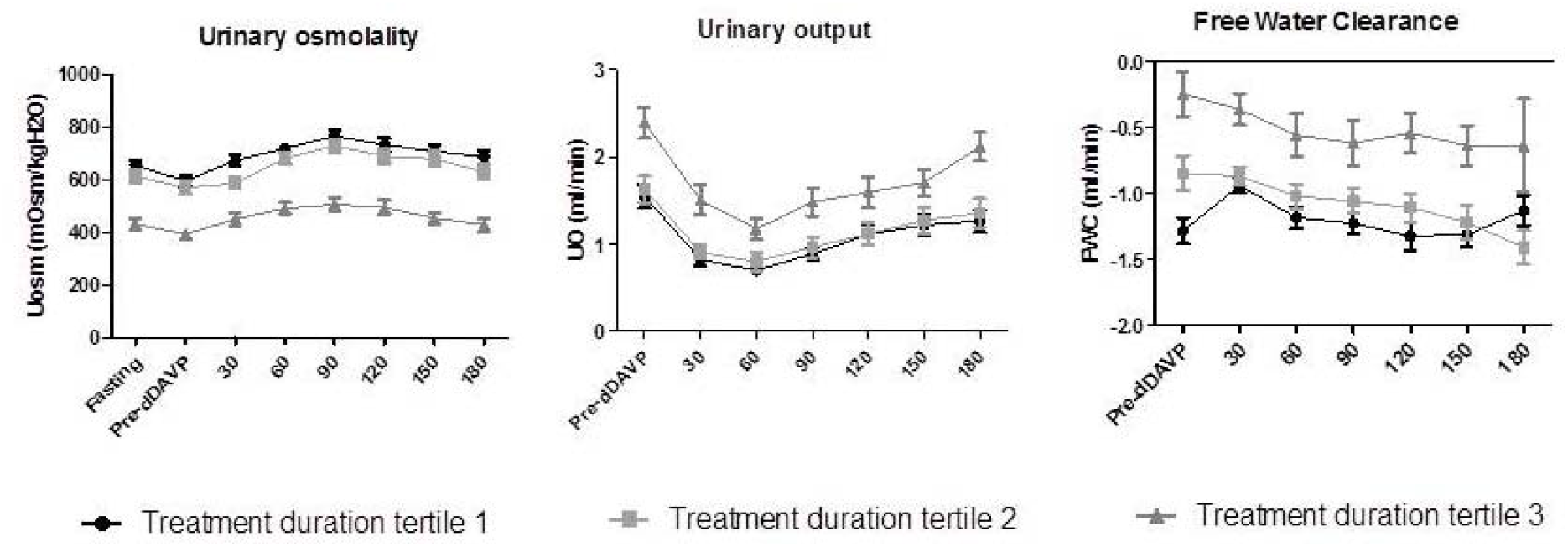
Water restriction and dDAVP test. Urinary osmolality, urinary output and free water clearance were assessed along the DDAVP test every 30 minutes. Tertile 1 of lithium treatment duration: <2.5 years; tertile 2 of lithium treatment duration: 2.5-10 years; tertile 3 of lithium treatment duration: >10 years.

### Individuals’ characteristics according to subgroups of urine concentrating features: impact of lithium daily dose on vasopressin levels and polyuria

The population was divided in 4 groups according to serum vasopressin levels and maximal urine concentrating ability as defined in the method section. This allowed differentiating between 1/ individuals with normal urine concentrating pattern (normal maximal urine osmolality and normal vasopressin level) (n=63) 2/ individuals with increased vasopressin level but still normal urine concentrating ability, suggesting early renal resistance to vasopressin (n=65) 3/ individuals with impaired urine concentrating ability and elevated vasopressin level (confirmed nephrogenic diabetes insipidus (n=54) 4/ individuals with decreased maximal urine osmolality but no elevation of vasopressin level (n=12) suggesting primary polydipsia (table 2). When comparing to group 1, individuals in the group 2 were significantly younger (p=0.03), with a significantly higher daily dose of lithium (p<0.001), higher plasma lithium concentrations (p=0.04), and higher copeptin levels (p<0.001) (table 2). There was no significant difference between the 2 groups in terms of lithium treatment duration. In the same line, the percentage of individuals with polyuria and with elevated vasopressin levels increased as lithium daily dose increased (figure 2), with 100% of individuals with a daily dose above 1400 mg/day displaying elevated vasopressin levels.

**Table 2.** Patients characteristics according to urine concentrating features.

|  | Group 1 | Group 2 |  | Group 3 | Group 4 |  |  |
| --- | --- | --- | --- | --- | --- | --- | --- |
|  | ADH<5 pg/ml<br>Max Uosm≥<br>600<br>mOsm/kgH<sub>2</sub>O | ADH≥5<br>pg/ml Max<br>Uosm ≥ 600<br>mOsm/<br>kgH<sub>2</sub>O | p-value<br>group 1<br>versus<br>group 2 | ADH≥5 pg/ml<br>Max Uosm <<br>600 mOsm/<br>kgH<sub>2</sub>O | ADH<5<br>pg/ml Max<br>Uosm < 600<br>mOsm/<br>kgH<sub>2</sub>O | p-value<br>group 3<br>versus<br>group 4 | p-value<br>all |
| n | 63 | 65 |  | 54 | 12 |  |  |
| Age, years | 50 [38- 63] | 40 [31- 58] | **0.03** | 60 [48- 66] | 51 [41- 57] | 0.05 | **<0.001** |
| Male sex n (%) | 25 (39.7) | 27 (41.5) | 1 | 15 (27.8) | 4 (33.3) | 1 | 0.4 |
| Lithium treatment<br>duration, years | 3 [2- 7] | 3 [1- 7] | 0.4 | 18 [10- 28] | 13 [4- 17] | 0.06 | **<0.001** |
| Sustained-release<br>formulation n (%) | 42 (68.9) | 50 (78.1) | 0.3 | 25 (46.3) | 8 (66.7) | 0.3 | **0.003** |
| Daily dose, mg/d | 800<br>[500- 1000] | 950<br>[750- 1200] | **<0.001** | 750<br>[500- 1000] | 800<br>[575- 1000] | 0.5 | **0.003** |
| Serum lithemia,<br>mmol/L | 0.65<br>[0.50- 0.85] | 0.77<br>[0.64- 0.87] | **0.04** | 0.72<br>[0.60- 0.89] | 0.76<br>[0.60- 0.88] | 0.8 | 0.2 |
| 24-hour urinary<br>ouput, mL/d | 1812<br>[1471- 2312] | 1558<br>[1337- 2060] | 0.08 | 2883<br>[2022- 3513] | 3026<br>[1755- 3786] | 1 | **<0.001** |
| Fasting thirst score,<br>0-10 scale | 6 [4- 7] | 6 [4- 8] | 0.6 | 5 [3- 7] | 5 [4- 5] | 0.1 | **0.04** |
| Fasting natremia,<br>mmol/L | 141<br>[140- 142] | 141<br>[140- 142] | 0.9 | 143<br>[141- 144] | 141<br>[141- 142] | 0.1 | **<0.001** |
| Maximal Uosm,<br>mOsm/kgH<sub>2</sub>O | 814<br>[709.50- 898] | 826<br>[739- 885] | 1 | 435<br>[318- 516] | 541<br>[483- 560] | **0.03** | **<0.001** |
| Plasma vasopressin,<br>pg/mL | 3.39<br>[2.90- 4.20] | 7.80<br>[5.80- 12.50] | **<0.001** | 10.55<br>[6.81- 16.73] | 3.69<br>[2.48- 4.38] | **<0.001** | **<0.001** |
| Plasma copeptin,<br>pmol/L | 8.12<br>[6.73- 10.89] | 13.30<br>[9.92- 19.25] | **<0.001** | 25.87<br>[18.30- 45.18] | 9.73<br>[3.04- 16.80] | **0.03** | **<0.001** |
| mGFR,<br>ml/min/1.73m<sup>2</sup> | 78.0<br>[72.4- 90.4] | 88.0<br>[76.8- 94.2] | 0.1 | 54.4<br>[43.1- 72.2] | 81.1<br>[58.6- 89.4] | **0.02** | **<0.001** |

Data are expressed as median (Q1-Q3) or number (percentage). Comparisons were performed using Mann-Whitney test when 2 groups were compared, and Kruskal-Wallis test when comparing all the groups for quantitative variables. Chi-squared test was performed for categorical variables. Uosm: urinary osmolality. mGFR: measured glomerular filtration rate.

**Figure 2.**
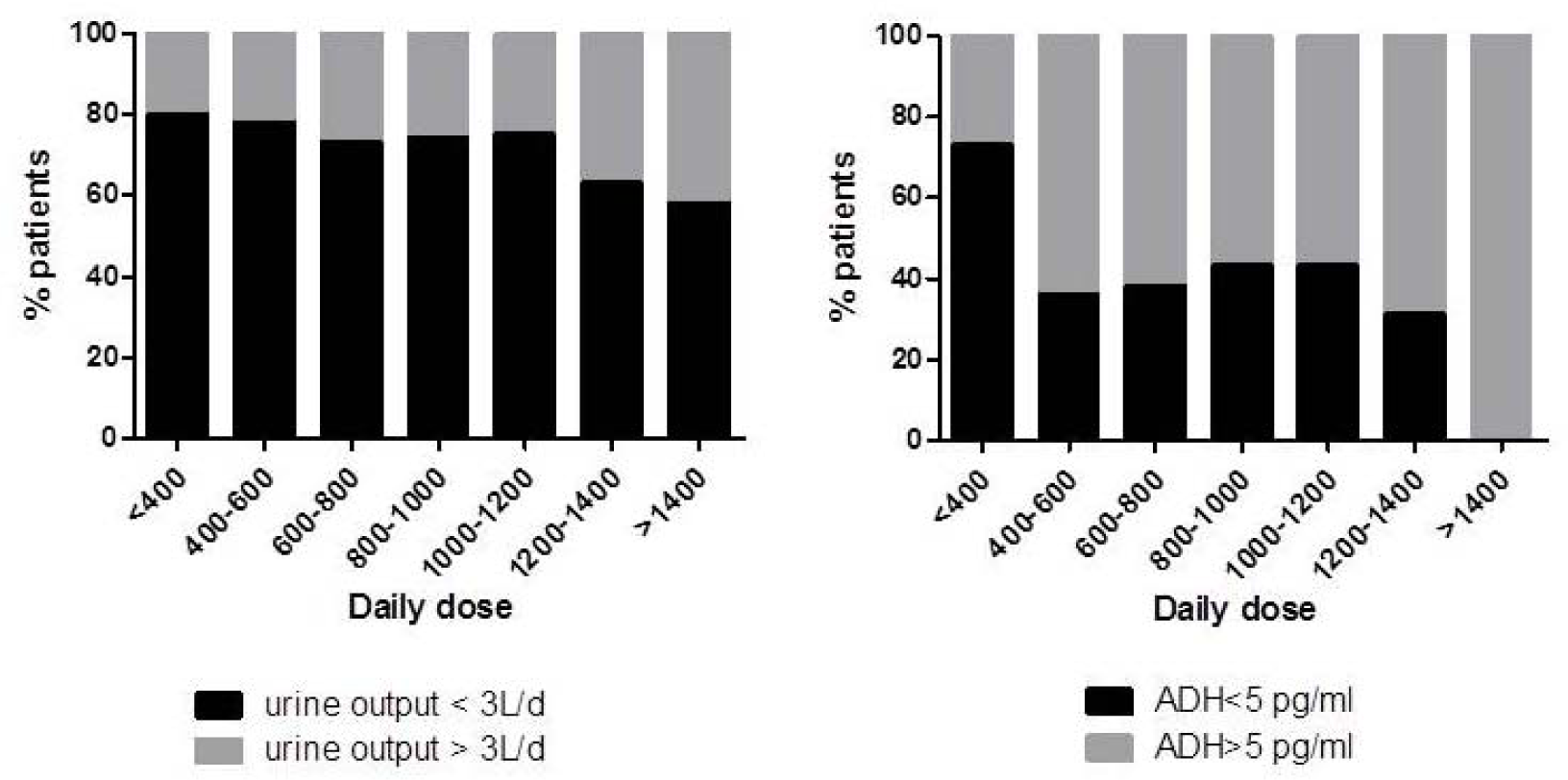
Polyuria and plasma vasopressin levels according to lithium daily dose. Daily dose is expressed in mg/day.

### Determinants of urinary output

When considering multivariable analysis, 24-hour urinary output was significantly and independently associated with a lower maximal urinary osmolality (β ± SE -2.89 ± 0.35, p<0.001), a higher osmolar intake (β ± SE 2.21 ± 0.24, p<0.001), a higher lithium daily dose (β ± SE 0.49 ± 0.17, p=0.005), and a higher vasopressin level (β ± SE 10.17 ± 4.76, p=0.03). It was also positively associated with female sex (β ± SE for male sex -358.74 ± 122.92, p=0.004). There was no association with lithium formulation (sustained-release or immediate-release). There was no association between lithium treatment duration and urine output when maximal urine osmolality was in the model, with no added value to the R-squared. Urine output was negatively associated with fasting thirst score (β ± SE -63.45 ± 23.61, p=0.008) (table 3).

**Table 3.**
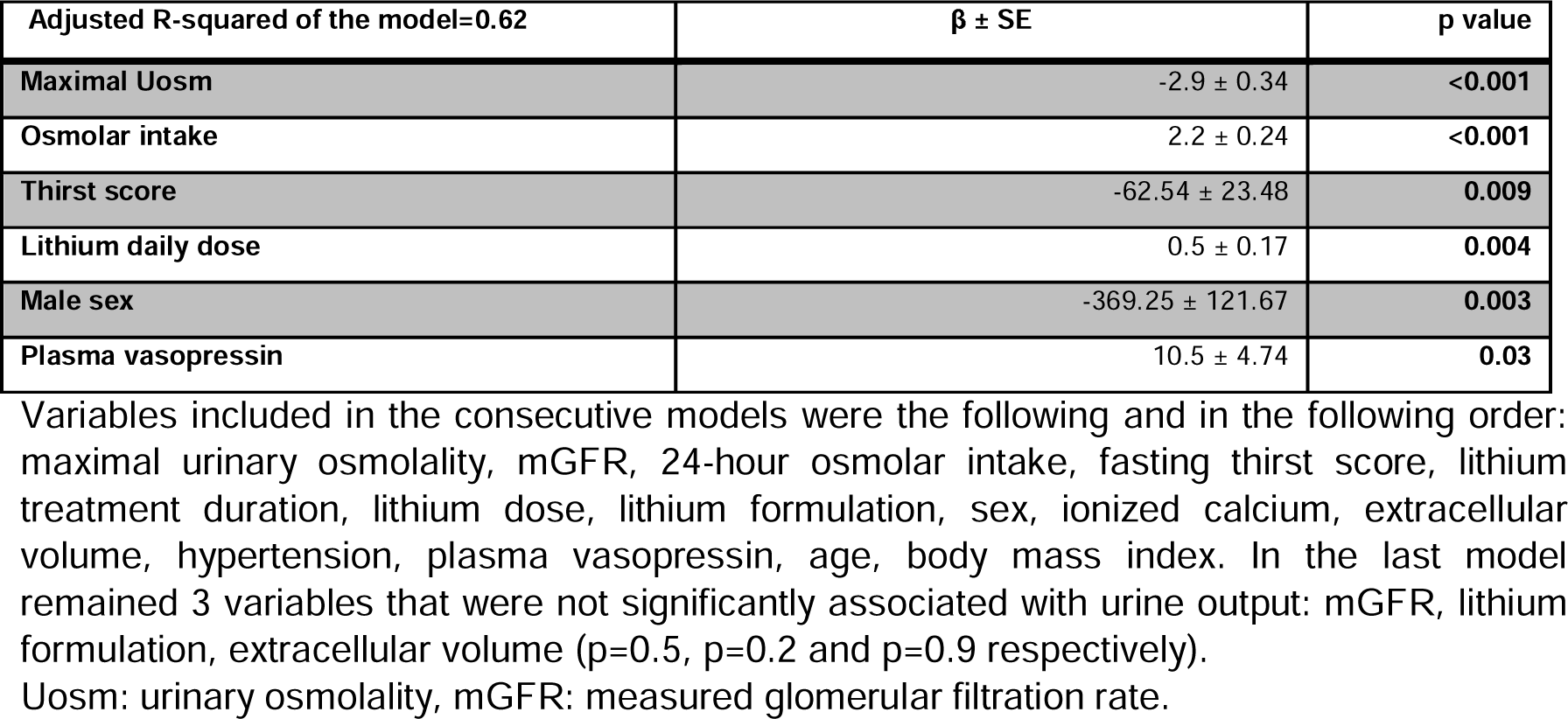
Multivariable analysis of the correlates of 24-hour urinary output.

| Adjusted R-squared of the model=0.62 | $\beta \pm SE$ | p value |
| --- | --- | --- |
| Maximal Uosm | -2.9 ± 0.34 | <0.001 |
| Osmolar intake | 2.2 ± 0.24 | <0.001 |
| Thirst score | -62.54 ± 23.48 | 0.009 |
| Lithium daily dose | 0.5 ± 0.17 | 0.004 |
| Male sex | -369.25 ± 121.67 | 0.003 |
| Plasma vasopressin | 10.5 ± 4.74 | 0.03 |

Variables included in the consecutive models were the following and in the following order: maximal urinary osmolality, mGFR, 24-hour osmolar intake, fasting thirst score, lithium treatment duration, lithium dose, lithium formulation, sex, ionized calcium, extracellular volume, hypertension, plasma vasopressin, age, body mass index. In the last model remained 3 variables that were not significantly associated with urine output: mGFR, lithium formulation, extracellular volume (p=0.5, p=0.2 and p=0.9 respectively).
Uosm: urinary osmolality, mGFR: measured glomerular filtration rate.

## Discussion

Our study conducted in a population of patients treated with lithium salts found a relatively low prevalence of polyuria (21%), but a majority of patients displayed a high concentration of serum vasopressin even with no overt concentrating ability defect. This was associated with a higher daily lithium dose. We also found that the major correlates of urine output in these patients were the daily osmolar intake, maximal urine osmolality, and lithium daily dose, independently of other factors such as treatment duration.

Although the exact prevalence remains unknown, lithium-induced polyuria has been reported to reach 20 to 70% depending on the studied populations, treatment duration and on the definition of polyuria, (5,9,19–22). In our population, the prevalence was relatively low. It is however important to mention that patients were systematically referred to the unit, whether they had symptoms or not.

Results of urine concentrating tests showed significant modifications of urine concentrating test features in patients belonging to the third tertile of treatment duration (*i*.*e*. > 10 years). On the contrary, no significant difference was observed between the two first tertiles of treatment duration.

More than half of the individuals had increased levels of serum vasopressin, a finding which was more frequent than overt polyuria. These levels were consistent with levels of serum copeptin in the subset of individuals for which they were available, accordingly with previously reported correlation between the two measurements (23). This suggests a certain degree of renal resistance to vasopressin, even in individuals not presenting yet a polyuria or altered urine concentrating ability. Another feature of vasopressin resistance was the low response to DDAVP test even though the majority of individuals did not reach hypernatremia at the time of DDAVP injection. Elevated levels of vasopressin have been reported in individuals with confirmed nephrogenic diabetes insipidus associated with lithium (24). This feature of near normal urine concentrating ability and elevated vasopressin or copeptin (the surrogate marker of vasopressin) levels has been reported in other conditions such as polycystic kidney disease (PKD) (25,26), and during chronic kidney disease (27), during which a decrease in urine concentrating ability is predictive of a steeper decline in mGFR and of ESKD (28). As it is known to play an important role in the pathophysiology of cyst growth in PKD and some authors suggest a deleterious role in CKD as well (29,30), one could hypothesize that it might also be an important mediator of lithium-induced tubulo-interstitial and microcystic kidney disease. This increase might also preclude overt NDI. In the same line, serum vasopressin was independently associated with a texture index that we recently developed based on kidney MRI radiomics analyses of a subset of the studied cohort (31).

Interestingly, while treatment duration affected urine concentrating ability after 10 years, we also found significantly lower doses of lithium in individuals treated more than 10 years. This result is related to the observational design of our study, as lithium dose might be lowered in patients with a stabilized disorder as recommended by good practice, or for a nephroprotective purpose. Interestingly, plasma lithium level was not correlated with treatment duration suggesting a dose adjustment to renal function.

Nevertheless, we found a significant relationship between urine output and lithium daily dose, independently of other factors including lithium treatment duration, as described by Kinahan *et al* (32). As expected, maximal urine osmolality was also a major determinant of urine output, which might be at least partly related to lithium treatment duration, explaining the absence of association between treatment duration and urinary output in the multivariable analysis (19). Daily osmolar intake, estimated by the 24-hour urine osmolality, was also a determinant of urine output, suggesting that lowering salt and protein intake in individuals with lithium-induced NDI may be advised to lower urine output in clinical practice (33). Female sex was associated with a higher urine output, consistent with a previous report by Schoen *et al*. in a population-based cohort of healthy subjects (34), but in contrast with a meta-analysis from Perucca *et al*. gathering studies in healthy subjects and in populations with CKD (35).

We evaluated the association between the galenic formulation of lithium therapy and diuresis and found no significant association. This is often a clinical practice issue when facing lithium-induced polyuria, as data from the literature are conflicting. Some authors have reported a lower risk of polyuria with the sustained-release formulation based on a low level of evidence (36–38), whereas others have hypothesized the contrary on the basis of a decreased renal parenchyma cumulative exposure time to lithium allowing renal cells to regenerate (39). Our study does not support any of these hypotheses, hence we could not recommend a specific formulation in order to prevent lithium-induced NDI.

We also analyzed thirst patterns in the population. We surprisingly found a negative association between fasting thirst score and diuresis, disproving the hypothesis of a central stimulation of thirst by lithium (40). In contrast, these results suggest that these individuals tend to adapt to the thirst distress in a certain degree.

It should be highlighted that the determinants of measured GFR were different from those of urinary output in our studied cohort (11). A functional effect of lithium on urine concentrating ability might thus result from various causes including daily dose, whereas lithium treatment duration, hence cumulative dose, was associated with a lower mGFR, suggesting two different mechanisms inducing these two renal side effects of lithium treatment.

Our study displays some limitations. Firstly, the cross-sectional and observational design of the study does not allow directly confirming the direct causal effect of lithium daily dose on 24-hour urinary volume. Secondly, missing data regarding plasma lithium levels prevented us from analyzing quantitative renal exposure to lithium. Thirdly, we did not perform a hyperosmotic test in order to test the spontaneous maximal urine osmolality before DDAVP injection. However, the occurrence of central diabetes insipidus is rare in this population (41) and DDAVP injection evidences the maximal urine osmolality. Moreover, our protocol was standardized in order to establish the same timepoints of DDAVP injection and urine and plasma measurements in all individuals.

In conclusion, our study suggests that lowering lithium daily dose might decrease 24-hour urine output, and that it should be considered in individuals affected by the disorder. Controlling salt and protein intake might also help decreasing 24-hour urine volume as it affects 24-hour urine osmolar output. We also found an early increase in vasopressin level in a majority of individuals probably even before the development of NDI, suggesting an early resistance to the action of vasopressin that might have potential deleterious role in the pathophysiology of CKD associated with chronic lithium treatment.

## Data Availability

All data produced in the present study are available upon reasonable request to the authors

## Acknowledgements

The authors wish to thank all the staff that was involved in the patients’ care and who accepted to be part of this study.

